# A Model for SARS-CoV-2 Infection with Treatment

**DOI:** 10.1101/2020.04.24.20077958

**Authors:** Amar Nath Chatterjee, Fahad Al Basir

**Affiliations:** Department of Mathematics, K.L.S. College, Nawada, Magadh University, Bodh Gaya, Bihar-805110, India; Department of Mathematics, Asansol Girls’ College, Asansol-4, West Bebgal-713304, India

**Keywords:** Mathematical model, Basic reproduction number, Stability analysis, Immunostimulant drug, Angiotensin Converting Enzyme, 2 (ACE2), Impulsive differential equation

## Abstract

The current emergence of coronavirus (SARS-CoV-2) puts the world in threat. The structural research on the receptor recognition by SARS-CoV-2 has identified the key interactions between SARS-CoV-2 spike protein and its host (epithelial cell) receptor, also known as angiotensin-converting enzyme 2 (ACE2). It controls both the crossspecies and human-to-human transmissions of SARS-CoV-2. In view of this, we propose and analyze a mathematical model for investigating the effect of CTL responses over the viral mutation to control the viral infection when a postinfection immunostimulant drug (pidotimod) is administered at regular intervals. Dynamics of the system with and without impulses have been analyzed using the basic reproduction number. This study shows that the proper dosing interval and drug dose both are important to eradicate the viral infection.

## 1 Introduction

A novel coronavirus named SARS-CoV-2 (an interim name proposed by WHO (World Health Organization) become pandemic since December 2019. The first infectious respiratory syndrome was recognised in Wuhan, Hubei province of China. Dedicated virologist identified and recognised the virus within a short time [3]. The SARS-CoV-2 is a single standard RNA virus genome which is closely related to severe acute respiratory syndrome SARS-CoV [4]. The infection of SARS-CoV-2 is associated with a SARS-CoV like a disease with a fatality rate of 3.4% [5]. The World Health Organisation (WHO) have named the disease as COVID-19 and declared it as a public health emergency worldwide [2].

The common symptoms of COVID-19 are fever, fatigue, dry cough, myalgia. Also, some patients suffer from headaches, abdominal pain, diarrhea, nausea, and vomiting. In the acute phase of infection, the disease may lead to respiratory failure which leads to death also. From clinical observation, within 1-2 days after patient symptoms, the patient becomes morbid after 4-6 days and the infection may clear within 18 days [6] depending on the immune system. Thus appropriate quarantine measure for minimum two weeks is taken by the public health authorities for inhibiting community spread [1].

In [3], Zhou *et al*. identified that the respiratory tract as principal infection site for COVID-19 infection. SARS-CoV-2 infects primary human airway epithelial cells. Angiotensic converting enzyme II (ACE2) receptor of epithelial cells plays an important role in cellular entry [3,8]. It has been observed that ACE2 could be expressed in the oral cavity. ACE2 receptors are higher in tongue than buccal and gingival tissues. These findings imply that the mucosa of the oral cavity may be a potentially high-risk route of COVID-19 infection. Thus epithelial cells of the tongue are the major routes of entry for COVID-19. Zhou et al. [3] also reported that SARS-CoV-2 spikes S bind with ACE2 receptor of epithelial cells with high affinity. The bonding between S - spike of SARS-CoV-2 with ACE2 [8], results from the fusion between the viral envelope and the target cell membrane and the epithelial cells become infected. The S protein plays a major role in the induction of protective immunity during the infection of SARS-CoV-2 by eliciting neutralization antibody and T cell responses [10]. S protein is not only capable of neutralizing antibody but it also contains several immunogenic T cell epitopes. Some of the epitopes found in either S1 or S2 domain. These proteins are useful for SARS-CoV-2 drug development [14].

We know that virus clearance after acute infection is associated with strong antibody responses. Antibody responses have the potential to control the infection [15]. Also, CTL responses help to resolve infection and virus persistence caused by weak CTL responses [9]. Antibody responses against SARS-CoV-2 play an important role in preventing the viral entry process [10]. Hsueh et al. [4] found that antibodies block viral entry by binding to the S glycoprotein of SARS-CoV-2. To fight against the pathogen SARS-CoV-2, the body requires SARS-CoV-2 specific *CD4+T* helper cells for developing this specific antibody [10]. Antibody-mediated immunity protection helps the anti-SARS-CoV serum to neutralize COVID-19 infection. Besides that, the role of T cell responses in COVID-19 infection is very much important. Cytotoxic T lymphocytes (CTLs) responses are important for recognizing and killing infected cells, particularly in the lungs [10]. But the kinetic of the CTL responses and antibody responses during SARS-CoV-2 infection is yet to be explored. Our study will focus on the role of CTL and its possible implication on treatment and drug development. the drug that stimulates the CTL responses represents the best hope for control of COVID-19. Here we have modeled the situation where CTLs can effectively control the viral infection when the post-infection drug is administered at regular intervals.

Mathematical modeling with real data can help in predicting the dynamics and control of an infectious disease [32,33]. A four-dimensional dynamical model for a viral infection is proposed by Tang et al. [11] for MERS-CoV mediated by DPP4 receptors. In case of SARS-CoV-2, the infection process is almost similar with MERS-CoV and SARS-CoV. For SARS-CoV-2 infection, the ACE2 receptor of epithelium cell are the major target area.

Since the dynamics of the disease transmission of SARS-CoV-2 in cellular level is yet to be explored, thus we investigate the system in the light of previous literature of [11,26-29] to formulate the dynamic model which play a significant role in describing the interaction between uninfected cells, free virus and CTL responses. We propose a novel deterministic model which describes the cell biological infection of SARS-CoV-2 with epithelial cells and the role of the ACE2 receptor.

We explained the dynamics in the acute infection stage. It has been observed that CTL proliferate and differentiate antibody production after they encounter antigen. Here we investigate the effect of CTL responses over the viral mutation to control viral infection when a post-infection drug is administered at regular intervals by mathematical perspective.

It is clinically evident that immunostimulants play a crucial role in the case of respiratory disease. Among the currently available immunostimulants, Pidotimod is the most effective for the respiratory disease [31]. Pidotimod increases the level of immunoglobulins (IgA, IgM, IgG) and activates the CTL responses to fight against the disease.

In this article, we have considered the infection dynamics of SARS-CoV-2 infection in the acute stage. We have used impulsive differential equations to study the immunostimulant drug dynamics and the effects of perfect drug adherence. In recent years the effects of perfect adherence have been studied by using impulsive differential equations in [12,13,17-21]. With the help of impulsive differential equations, the effect of maximal acceptable drug holidays and optimal dosage can be found more precisely [12,21].

The article is organised as follows: The very next section contains the formulation of the impulsive mathematical model. Dynamics of the system without impulses has been provided in section 3. The system with impulses has been analysed in section 4. Numerical simulations, on the basis of the outcomes of section 3 and 4, have been included in section 5. Discussion in section 6 concludes the paper.

## 2 Model formulation

As discussed in the previous section, we propose a model considering the interaction between epithelium cells and SARS-CoV-2 virus along with lytic CTL responses over the infected cells. We consider five populations namely the uninfected epithelium cells *T*(*t*), infected cells *I*(*t*), ACE2 receptor of the epithelial cells *E*(*t*) and CTLs against the pathogen *C*(*t*).

lIn this model, we consider which represents the concentration of ACE2 on the surface of uninfected cells, which can be recognized by surface spike (S) protein of SARS-CoV-2 [24].

It is assumed that the susceptible cells are produced at a rate λ_1_ from the precursor cells and die at a rate *d_T_*. The susceptible cells become infected a rate *βE*(*t*)*V*(*t*)*T*(*t*). The constant *d_I_* is the death rate of the infected cells. Infected cells are also cleared by the body’s defensive CTLs at a rate p.

The infected cells produce new viruses at the rate *md_I_* during their life, and *d_V_* is the death rate of new virions, where *m* is any positive integer. It is also assumed that ACE2 is produced from the surface of uninfected cells at the constant rate λ_2_ and the ACE2 is destroyed, when free viruses try to infect uninfected cells, at the rate *θβE*(*t*)*V*(*t*)*T*(*t*) and is hydrolyzed at the rate *d_E_E*.

CTL proliferation in the presence of infected cells is described by the term

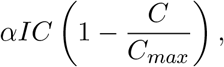

which shows the antigen dependent proliferation. Here we consider the logistic growth of CTL with *C_max_* as the maximum concentration of CTL and *d_c_* is its rate of decay.

With the above assumptions, we have the following mathematical model characterising the SARS-CoV-2 dynamics:

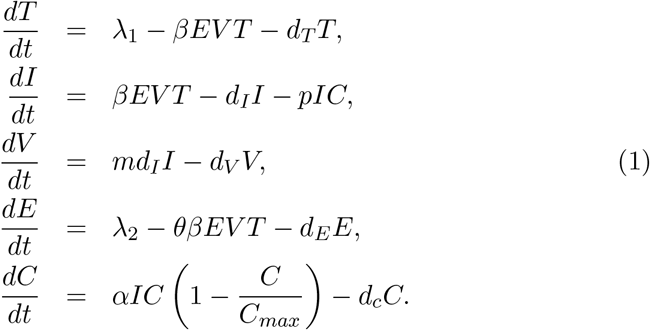

We now modify the above model incorporating pulse periodic drug dosing using impulsive differential equation [22,23].

We consider the perfect adherence behaviour of immunostimulant drug for SARS-CoV-2 infected patients at fixed drug dosing times *t_k_, k* ∊ ℕ.

We assume that CTL cells increases by a fixed amount *ω*, which is proportional to the total number of CTLs that the drug can stimulate. Thus the above model takes the following form:

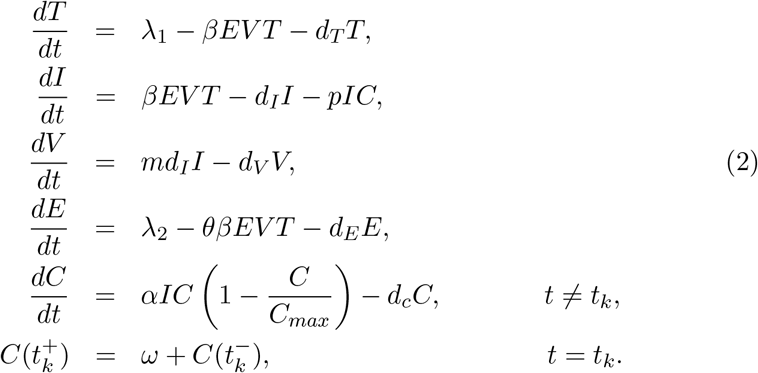

Here, 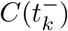 denotes the CTL cells concentration immediately before the impulse, 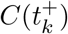 denotes the concentration after the impulse and *ω* is the fixed amount which is proportional to the total number of CTLs the drug stimulates at each impulse time *t_k_, k* ∊ ℕ.

Remark 1.

*It can be noted that when there is no drug application in the system, model* (2) *becomes model* (1).

## 3 Analysis of the system without drug

In this section, we analyse the dynamics of the system without impulses i.e. system (1). We have derived the *basic reproduction number* for the system. Stability of equilibria are discussed using the number.

### 3.1 Existence of equilibria

Model (2) has three steady states namely (i) the disease-free equilibrium 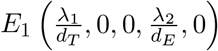, (ii) with 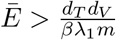, there is a CTL-responses-free equilibrium, 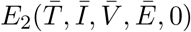, where,

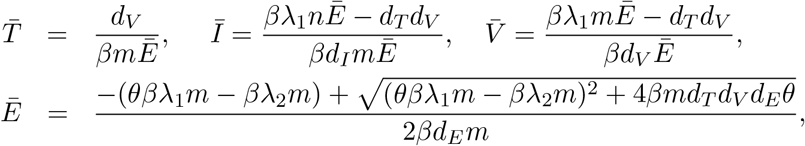

and (iii) the endemic equilibrium *E*^*^ which is given by

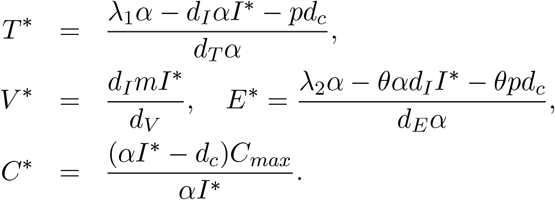

where, *I*^*^ is the positive root of the cubic equation

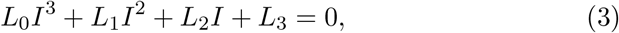

with,

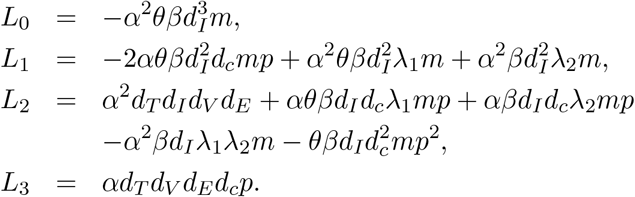

#### Remark 2.

*Note that L_0_ <* 0 *and L_3_ >* 0. *Thus, the equation* (3) *has at least one positive real root. If L_1_ >* 0 *and L_2_ <* 0, *then* (3) *can have two positive roots. For a feasible endemic equilibrium we also need*

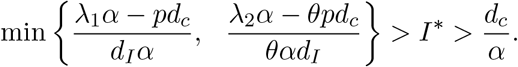

### 3.2 Stability of equilibria

In this section, the characteristic equation at any equilibria is determined for the local stability of the system (2). Linearizing the system (2) at any equilibria *E*(*T, I, V, E, C*) yields the characteristic equation

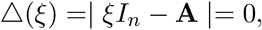

where *I_n_* is the identity matrix and **A** = [*a_ij_*] is the following 5 × 5 matrix given by

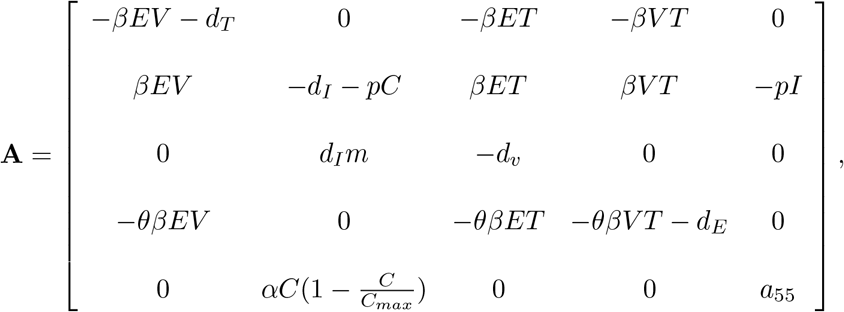

with 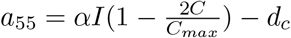. We finally get the characteristic equation as

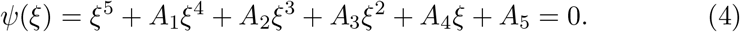

The coefficients *A_i_, i* = 1, 2,…, 5 are given in **Appendix-A**.

Looking at stability of any equilibrium *E*, the Routh-Hurwitz criterion gives that all roots of this characteristic equation (4) have negative real parts, provided the following conditions hold

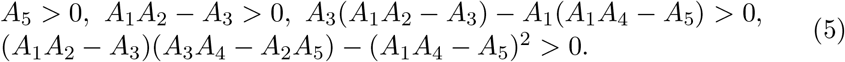

et us define the *basic reproduction number* as

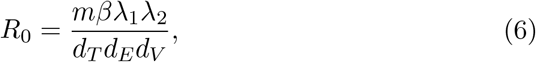

then using (5) we can derived the following result:

#### Theorem 1.

*Disease-free equilibrium*, 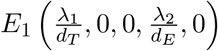 *of the model* (*2*) *is stable for R*_0_ < 1, *and unstable for R*_0_ > 1.

At *E*_2_ one eigenvalue is −*d_c_* and rest of the eigenvalues satisfy the following equation

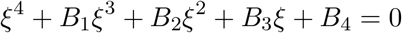

The coefficients *B_i_, i* = 1, 2,…,5 are given in **Appendix-B**.

Using Routh-Hurwitz criterion, we have the following theorem.

#### Theorem 2.

*CTL-free equilibrium*, 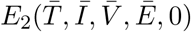, *is asymptotically stable if and only if the following conditions are satisfied*

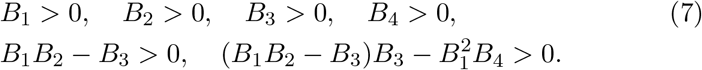

Denoting 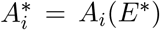 and using (5), we have the following theorem establishing the stability of coexisting equilibrium *E*^*^.

#### Theorem 3.

*The coexisting equilibrium E^*^ is asymptotically stable if and only if the following conditions are satisfied*

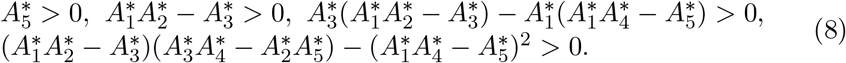

## 4 Dynamics of the system with impulsive drug dosing

In this section we consider the model system (2). Before analysing the system, we first discuss the one dimensional impulse system as follows:

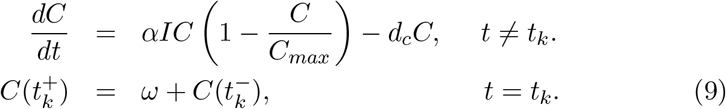

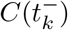 denotes the CTL responses immediately before the impulse drug dosing, 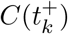 denotes the concentration after the impulse and *ω* is the dose that is taken at each impulse time *t_k_*, *k ∊* ℕ.

We now consider the following linear system,

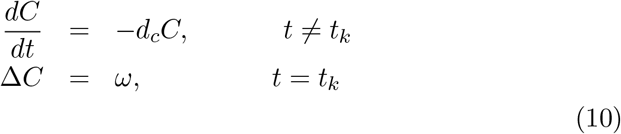

where, 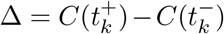. Let *τ* = *t_k_*_+1_ − *t_k_* be the period of the campaign. The solution of the system (10) is,

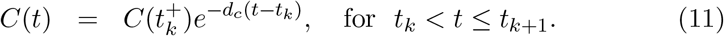

In presence of impulsive dosing, we can get the recursion relation at the moments of impulse as,

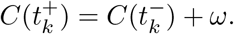

Thus the amount of CTL before and after the impulse is obtained as,

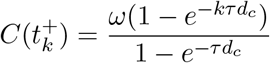

and

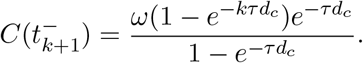

Thus the limiting case of the CTL amount before and after one cycle is as follows:

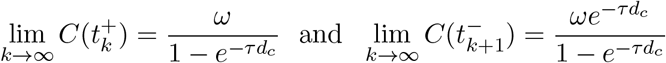

and

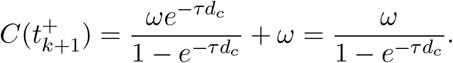

### Definition 1.

*Let* Λ ≡ (*S_u_, S_a_, I, C*), 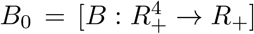, *then we say that B belong to class B*_0_ *if the following conditions hold:*

(*i*) *B is continuous on* 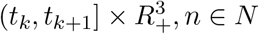 *and for all* Λ ∊ *R*^4^, 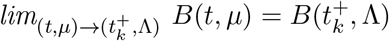 *exists*,

(*ii*) *B is locally Lipschitzian in* Λ.

We now recall some results for our analysis from [22,23].

Lemma 1.

*Let Z*(*t*) *be a solution of the system* (*9*) *with Z*(0^+^) ≥ 0. *Then Z_i_*(*t*) ≥ 0, *i* = 1,…, 4 *for all t ≥* 0. *Coreover, Z_i_*(*t*) *>* 0, *i* = 1,…, 4 *for all t >* 0 *if Z_i_*(0^+^) > 0, *i* = 1,…, 4.

Lemma 2.

*There exists a constant γ such that T*(*t*) ≤ γ, *I*(*t*) ≤ γ, *V*(*t*) ≤ *γ E*(*t*) ≤ *γ and C*(*t*) ≤ *γ for each and every solution Z*(*t*) *of system* (*9*) *for all sufficiently large t*.

Lemma 3.

*Let B ∊ B_0_ and also consider that*

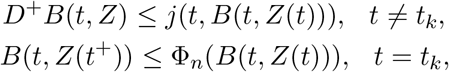

*where j*: **R**_+_ × **R**_+_ → **R** *is continuous in* (*t_k_,t_k+_*_1_] *for* 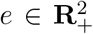, *n ∊ N, the limit* 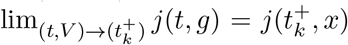 *exists and* 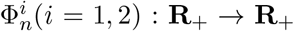 *is non-decreasing. Let y*(*t*) *be a maximal solution of the following impulsive differential equation*

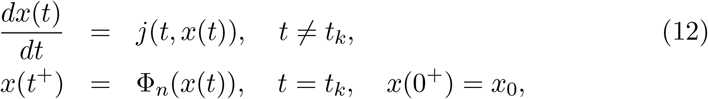

*existing on* (0^+^, ∞). *Then B*(0*^+^, Z*_0_) *< x_0_ implies that B*(*t, Z*(*t*)) ≤ *y*(*t*)*, t ≥* 0, *for any solution Z*(*t*) *of system* (*9*)*. If j satisfies additional smoothness conditions to ensure the existence and uniqueness of solutions for* (*12*)*, then y*(*t*) *is the unique solution of* (*12*).

We now consider the following sub-system:

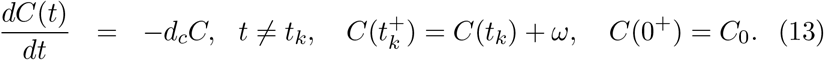

The Lemma provided above, gives the following result,

Lemma 4.

*System* (*13*) *has a unique positive periodic solution* 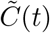 *with period τ and given by*

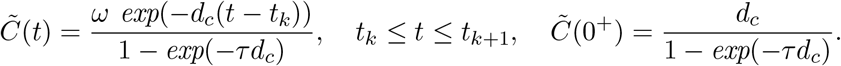

We use this result to derive the following theorem.

Theorem 4.

*The disease-free periodic orbit* (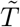, 0, 0, 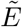, 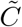) *of the system* (*2*) *is locally asymptotically stable if*

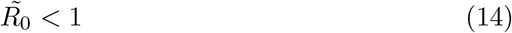

*where*,

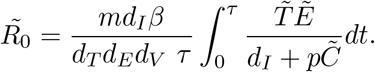

Proof.

Let the solution of the system (9) without infected people be denoted by (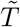, 0, 0, 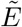, 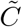), where

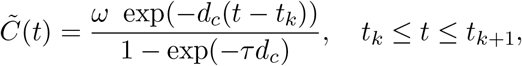

with initial condition *C*(0^+^) as in Lemma 4. We now test the stability of the equilibria. The variational matrix at (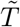, 0,0, 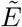, 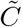) is given by

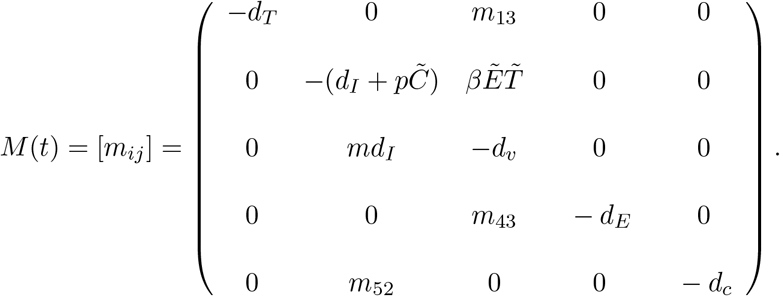

The monodromy matrix P of the variational matrix *M*(*t*) is

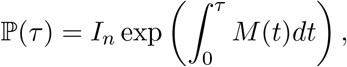

where *I_n_* is the identity matrix. Note that *m*_13_*,m*_43_*,m*_52_ are not required for this analysis, therefore we have not mentioned their expressions.

We can write 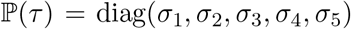, where, *σ_i,_ i* = 1, 2, 3, 4, 5, are the Floquet multipliers and they are determined as

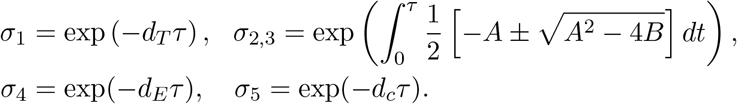

Here 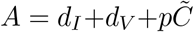 and 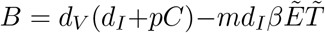. Clearly λ_1,4,5_ < 1. It is easy to check that *A*^2^ − 4*B >* 0 and if *B* ≥ 0 and hold then we have *λ*_2_, _3_ < 1. Thus, according to Floquet theory, the periodic solution (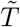, 0,0, 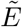, 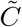) of the system (9) is locally asymptotically stable if the conditions given in (14) hold. □

**Table 1:**
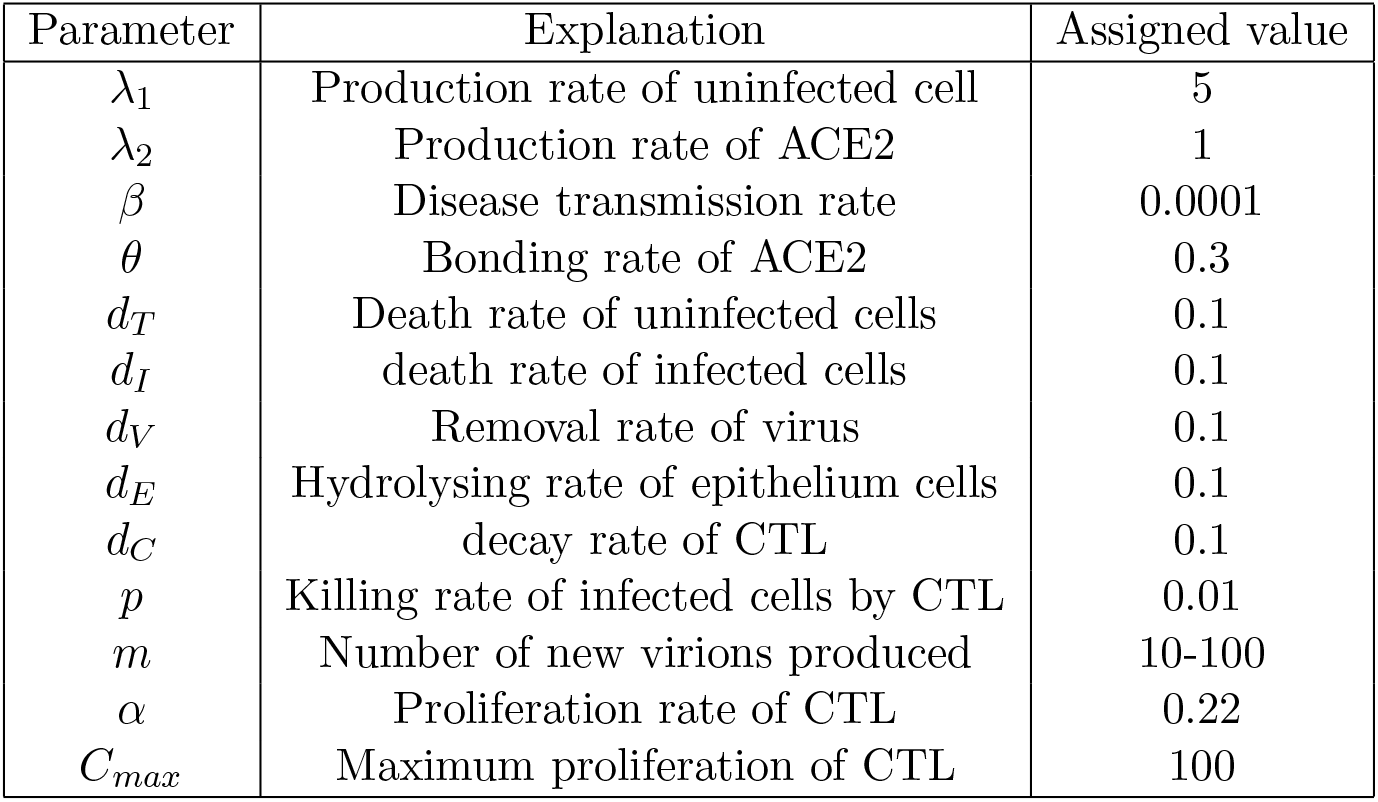
Set of parameter values used of numerical simulations.

| Parameter | Explanation | Assigned value |
| --- | --- | --- |
| $\lambda_1$ | Production rate of uninfected cell | 5 |
| $\lambda_2$ | Production rate of ACE2 | 1 |
| $\beta$ | Disease transmission rate | 0.0001 |
| $\theta$ | Bonding rate of ACE2 | 0.3 |
| $d_T$ | Death rate of uninfected cells | 0.1 |
| $d_I$ | death rate of infected cells | 0.1 |
| $d_V$ | Removal rate of virus | 0.1 |
| $d_E$ | Hydrolysing rate of epithelium cells | 0.1 |
| $d_C$ | decay rate of CTL | 0.1 |
| $p$ | Killing rate of infected cells by CTL | 0.01 |
| $m$ | Number of new virions produced | 10-100 |
| $\alpha$ | Proliferation rate of CTL | 0.22 |
| $C_{max}$ | Maximum proliferation of CTL | 100 |

## 5 Numerical results and discussion

In this section, we have observed the dynamical behaviours of system without drug (Figure 1 and Figure 2) and with impulsive effect of the drug dose (Figure 3 and Figure 4) through numerical simulations taking the parameters mainly from [11,30,31].

**Figure 1:**
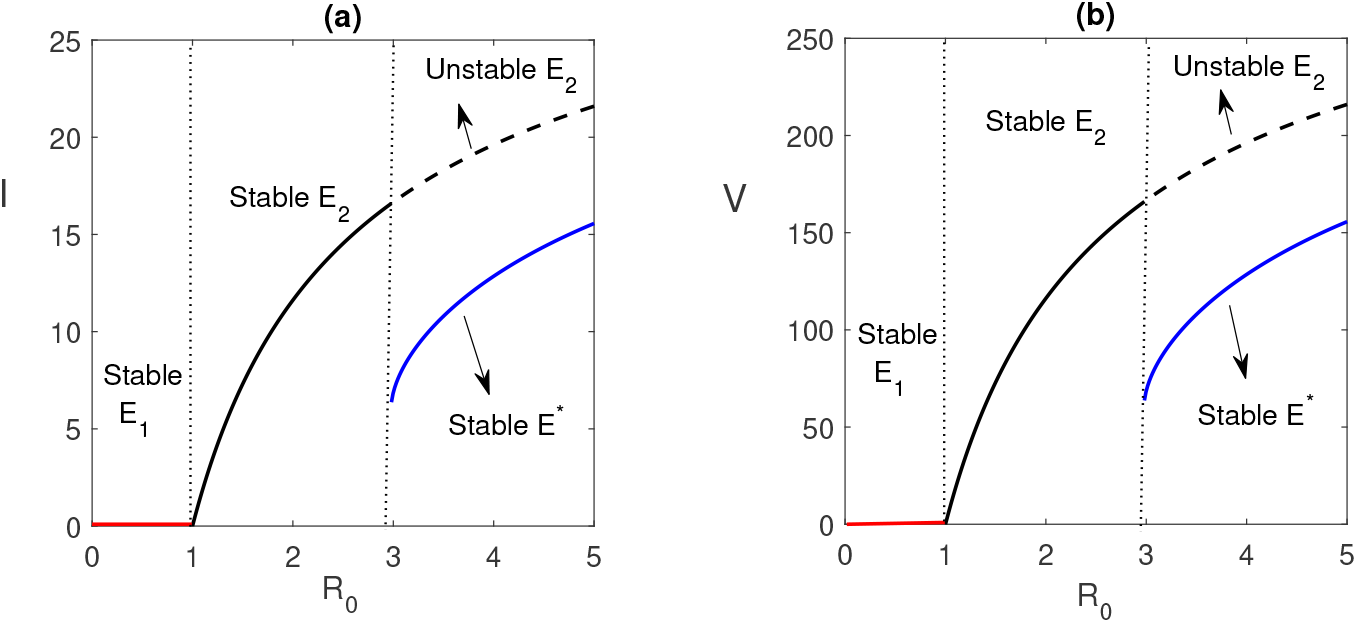
Existence and stability of equilibria is shown with respect to R0. Parameters values used in this figure are taken from Table 1 and *m* = 10. We have varied the value of *β* in (0.00001, 0.0001).

**Figure 2:**
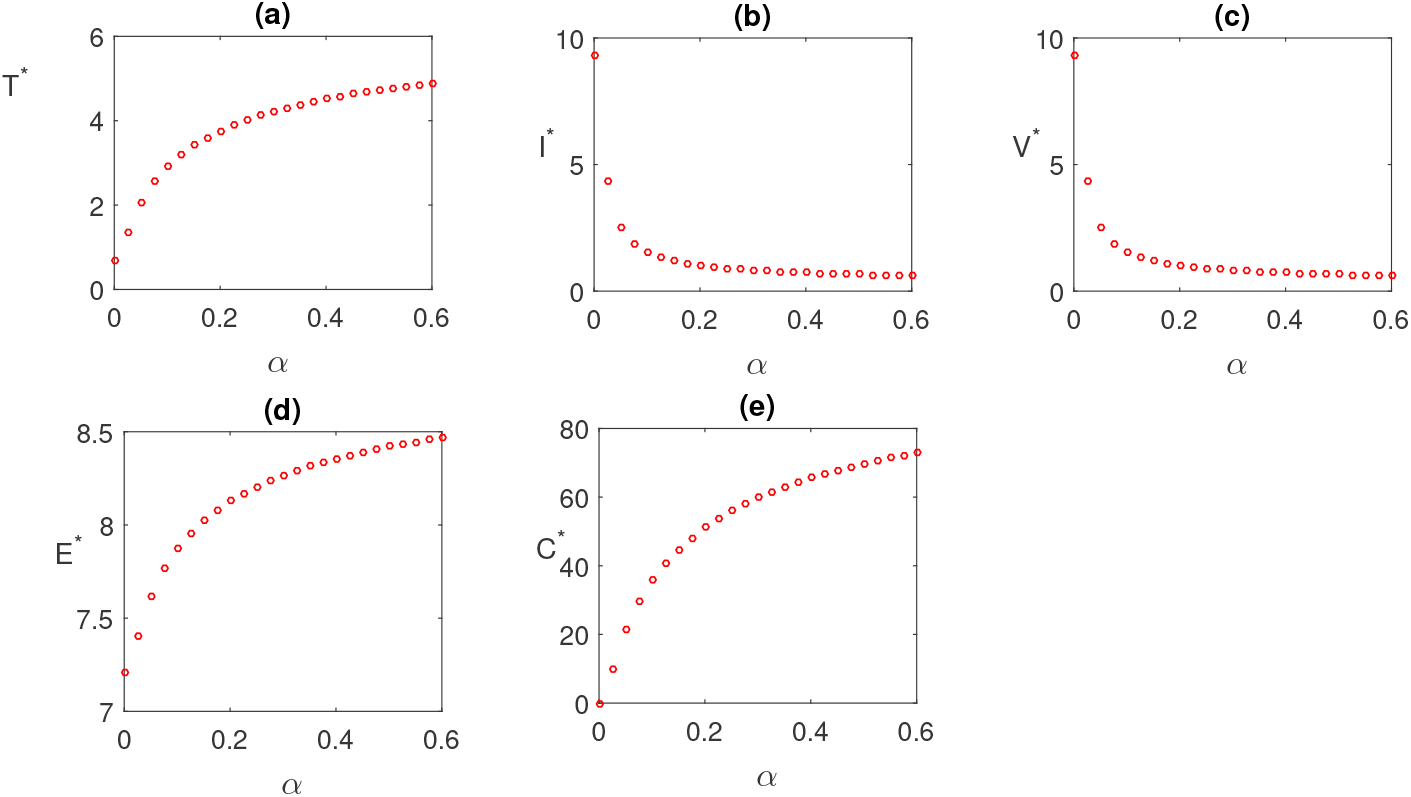
In the absence of the drug, effect of growth rate of CTL i.e. *α* on the steady state values of model population is shown. Parameters values used in this figure are same as Figure 1 except *α*.

We have mainly focused on the role of CTL and its possible implication on the treatment and drug development. The drug that stimulates the CTL responses represents the best hope for control of COVID-19. Here we have determined the situation where CTLs can effectively control the viral infection when the post-infection drug is administered at regular intervals.

Existence of equilibria of the system without drug dose is shown for different values of basic reproduction number *R*_0_. In plotting the Figure 1, we have varied the value of infection rate *β*. It is observed that for lower infection rate (that corresponds to *R*_0_ < 1) disease free equilibrium *E*_1_ is stable (corroborated with Theorem 1). It becomes unstable and ensure the existence of CTL-free equilibrium *E*_2_ which is stable if *R*_0_ < 2.957 (which corresponds to *β* = 0.00005963) and unstable otherwise. (This satisfies Theorem 2). Again we see that when *E*_2_ is unstable the *E^*^* is feasible. Also whenever *E^*^* exists, it is stable which verified the Theorem 3.

Effect of immune response rate *α* is plotted in Figure 2. We observe that in the absence of Drug, the CTL count and ACE2 increases with increasing value of *α*. Steady state value of infected cell *I\** and virus *V\** decreases significantly as *α* increases.

Due to the impulsive nature of the drugs, there are no equilibria of the system i.e. population do not reach to towards equilibrium point, rather approach a periodic orbit. Hence, we evaluate equilibrium-like periodic orbits. There are two periodic orbits of the system (2) namely the disease-free periodic orbit and endemic periodic orbit. Here our aim is to find the stability of disease-free periodic orbit.

Figure 3 compare the system without and with impulse Drug effect. In the absence of Drug we observe that the CTL count approaches a stable equilibrium. Under regular drug dosing, the CTL count oscillates in an impulsive periodic orbit. Assuming perfect adherence, if the drug is sufficiently strong, both infected cell and virus population are approaches towards extinction. In this case, the total number of uninfected cells reach its maximum level which implies that the system approaches towards its infection free state (Theorem 4).

**Figure 3:**
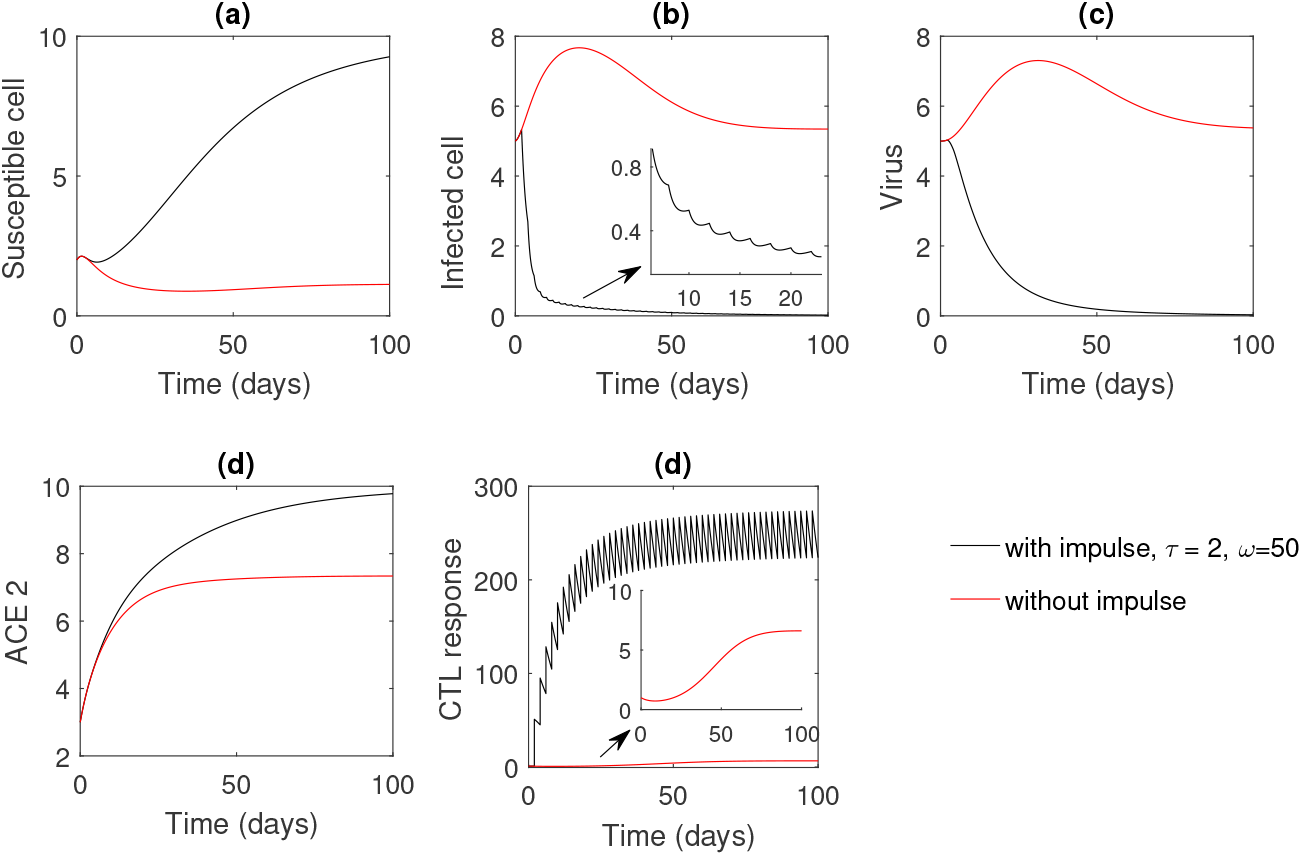
Numerical solution of the model system with and without drug dose is shown taking parameters as in Figure 1. In this figure, *τ* = 2, *ω* = 50.

If we take sufficiently large impulsive interval *τ* = 5 days (keeping rate *ω* = 50 fixed, as in Figure 3) or lower dosage effect *ω* = 20 (keeping interval *τ* = 2 fixed, as in Figure 3), in both the cases, infection is remains present in the system. Thus proper dosage of drug and optimal dosing interval are important for infection management.

## 6 Conclusion

In this article, the role of immunostimulant drug (mainly Pidotimod) during interactions between SARS-CoV-2 spike protein and epithelial cell receptor ACE2 in COVID-19 infection has been studied as a possible drug dosing policy. To reactivate the CTL responses during the acute infection period, immune activator drugs is delivered to the host system in an impulsive mode.

The immunostimulant drug when administered, the best possible CTL responses can act against the infected or virus-producing cells to neutralize infection. This particular situation can keep the infected cell population at a very low level. In the proposed mathematical model, we have analyzed the optimal dosing regimen for which infection can be controlled.

From this study, it has been observed that when the basic reproduction ratio lies below one, we expect the system to attain its disease-free state. However, the system switches from disease-free state to CTL-free equilibrium state when 1 *< R*_0_ < 2.957. If *R*_0_ > 2.957, the CTL-free equilibrium moves to an endemic state (Figure 1).

Here we have explored the immunostimulant drug dynamics by the help of impulsive differential equations. With the help of impulsive differential equations, we have studied how the effect of maximal acceptable optimal dosage can be found more precisely. Impulsive system shows that proper dosage and dosing intervals are important for the eradication of the infected cells and virus population which results the control of pandemic (Figure 3).

It has also been observed that the length of the dosing interval and the drug dose play a very decisive role to control and eradicate the infection. The most interesting prediction of this model is that effective therapy can often be achieved, even for low adherence, if the dosing regimen is adjusted appropriately (Figure 4). Also if the treatment regimen is not adjusted properly, the therapy is not effective at all. This approach might also be applicable to a combination of antiviral therapy.

**Figure 4:**
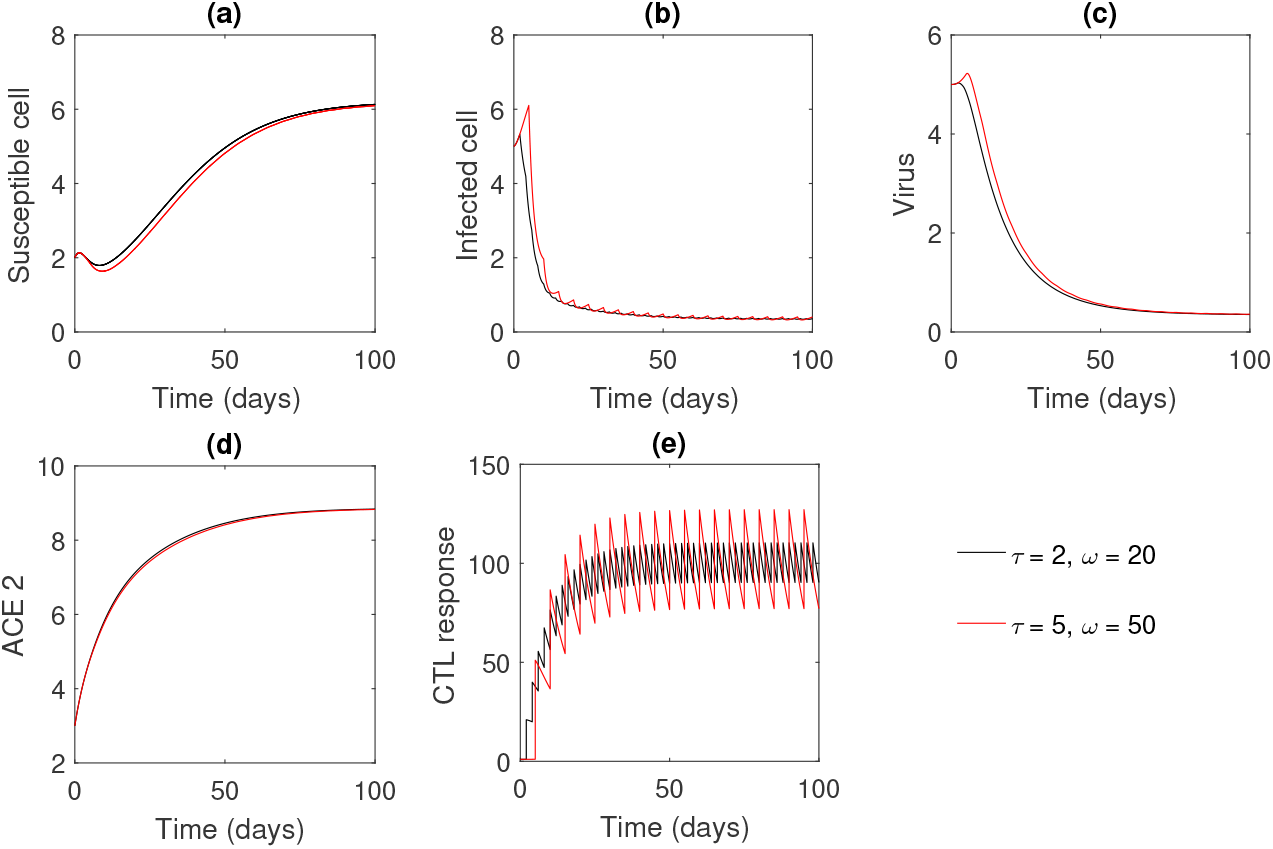
Numerical solution of the model system for different rates of Drug dosing and different intervals of impulses.

Future extension work of the combination of drug therapy should also include more realistic patterns of non-adherence (random drug holidays, imperfect timing of successive doses), more accurate intracellular pharmacokinetics and leads towards better estimates of drug dosage and drug dosing intervals.

We end the paper with the quotation: *“This outbreak is a test of political, financial and scientific solidarity for the world to fight a common enemy that does not respect borders…, what matters now is stopping the outbreak and saving lives.”* by Dr. Tedros, Director General, WHO [25].

## Data Availability

The methods and results data used to support the findings of this study are included in the article.

## Data Availability

The data used for supporting the findings are included within the article.

## Authors Contributions

Both authors contributed equally to this work.

## Appendix-A

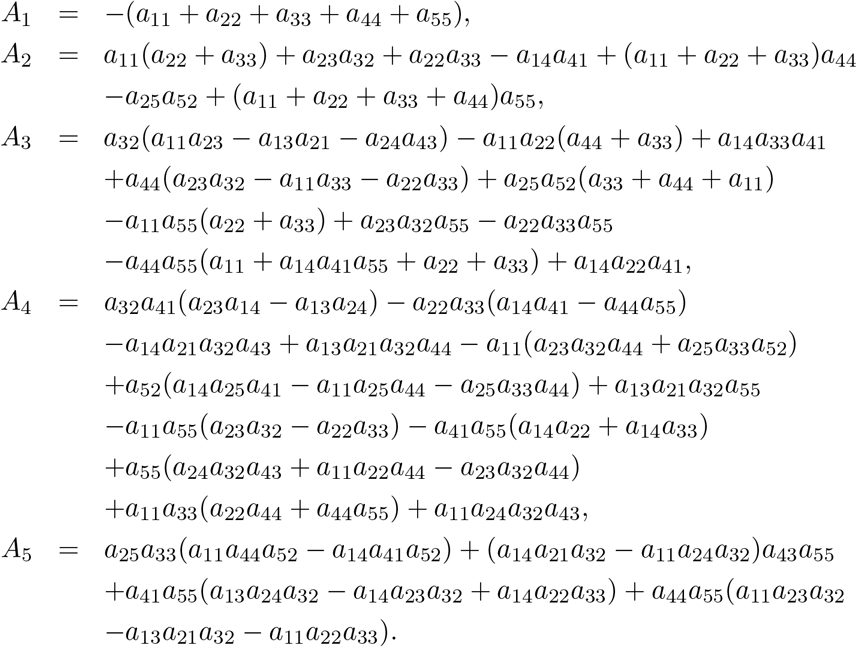

## Appendix-B

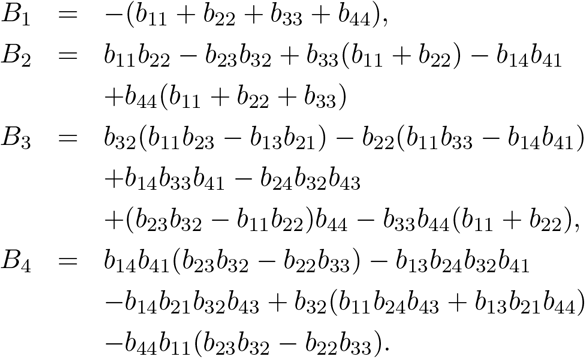

